# LABOR DURATION IS DEFINED BY THE TIME OF DAY OF LABOR INDUCTION

**DOI:** 10.1101/2024.10.14.24315464

**Authors:** Kylie Cataldo, Robert Long, Isoken Olomnu, Rene Cortese, Hanne M. Hoffmann

## Abstract

**BACKGROUND:** Spontaneous labor and birth peak during the late evening and early morning hours, indicating an endogenous rhythm in labor onset and birth. We hypothesize that the time-of-day of labor induction will define labor duration and the risk of cesarian section.

**METHODS:** In a retrospective study of pregnant women who were induced for labor (n =3,688), time-of-day of labor induction was studied across maternal phenotypes. Survival analysis and Cox Proportional Hazards model were used to identify differences in time-to-birth as a result of inducing labor at a specific time-of-day.

**RESULTS:** Labor induction was circadian (p<0.05, Lomb-Scargle test), with a gradual lengthening in labor duration when labor induction was initiated later in the day, peaking at 23:00 hours (average labor duration of 20.72 hours) as compared to induction at 5:00 hours (average labor duration of 14.74 hours, p<0.01, Kruskal-Wallis test). The optimal time-of-day of labor induction was conditioned by maternal phenotype with significant differences in probability of giving birth as a result of the time-of-day labor was induced for nulliparous obese (p<0.05, Two-way ANOVA), and parous obese women (p<0.05).

**CONCLUSIONS:** Labor duration in response to induction is circadian, with the shortest labor duration when induced during early morning hours. The optimal time-of-day of labor induction is conditioned by maternal phenotype and should be considered as a labor management practice.

## INTRODUCTION

Labor induction is an increasingly common obstetric intervention for initiating the onset of labor. The induction of labor is indicated when maternal and fetal outcomes are expected to improve with management as opposed to waiting for spontaneous labor to occur^1^. Common indications for labor induction include risk reduction after 39 weeks^2^ and complications such as carrying the fetus past term^3^, premature rupture of membranes^4^, health conditions such as gestational diabetes^5^ or preeclampsia^6^, and infection.

Synthetic oxytocin, Pitocin, is used to artificially induce labor by stimulating uterine contractions, mimicking the role of oxytocin during spontaneous labor^7^. It has been well established that spontaneous labor and birth peak during late evening and early morning hours^8,9^. This daily peak in birthrate indicates that circadian mechanisms contribute to labor onset. The contribution of the circadian system to labor onset is supported by the mouse uterus presenting with circadian rhythms^10^, oxytocin receptor knock-out mice lacking a circadian gating of labor onset^11^, and both mice and humans have a circadian release of oxytocin. In addition, we^12–15^ and others^16^ have identified that women with pregnancy complications, including preterm birth, preeclampsia and gestational diabetes, have deregulated expression of molecular clock genes which drive circadian rhythms on a cellular level, indicating that a deregulation of the bodies circadian timekeeping system is associated with poor pregnancy outcomes.

Here, we describe for the first time an important relationship between the time-of-day of labor induction and labor duration. We identify a significant decrease in average labor duration for women induced during early morning hours. Importantly, we identify that the association between labor duration and the time-of-day of labor induction (TOI) is related to BMI and parity, and shorter labor duration reduces the rate of cesarean section.

## METHODS

### Participants

This project is approved by the Institutional Review Board of the Michigan State University under Study ID: 0007199. Deidentified data was collected from Electronic Medical Records corresponding to deliveries (n=7,957) at the Sparrow Health System, East Lansing, MI from February 2019 to March 2022. After excluding subjects with incomplete data (n=4,269), differences in TOI were studied for women in whom labor was induced (n=3,688). Table 1 summarizes the demographic and clinical characteristics of the study population. Labor duration after induction was calculated by subtracting TOI from time of delivery. TOI was defined by the time of the initial dose of cervical ripening agent or Pitocin, whichever was first. Body Mass Index (BMI) was grouped according to the Centers for Disease Control and Prevention^17^ with BMI between 18.5 to 24.9 as normal, 25 to 29.9 as overweight, and greater than 30 as obese. There were no underweight subjects. Term deliveries were grouped according to gestational age (GA) with early term deliveries between 37 weeks 0 days and 38 weeks 6 days, term deliveries between 39 weeks 0 days and 40 weeks 6 days, and late term deliveries between 41 weeks 0 days and 41 weeks 6 days. NICU admission also included neonates admitted under NICU observation as Sparrow Hospital does not have a separate newborn nursery. Parity was defined at time of admission with P0 grouped as nulliparous and P1+ grouped as parous.

**Table 1.**
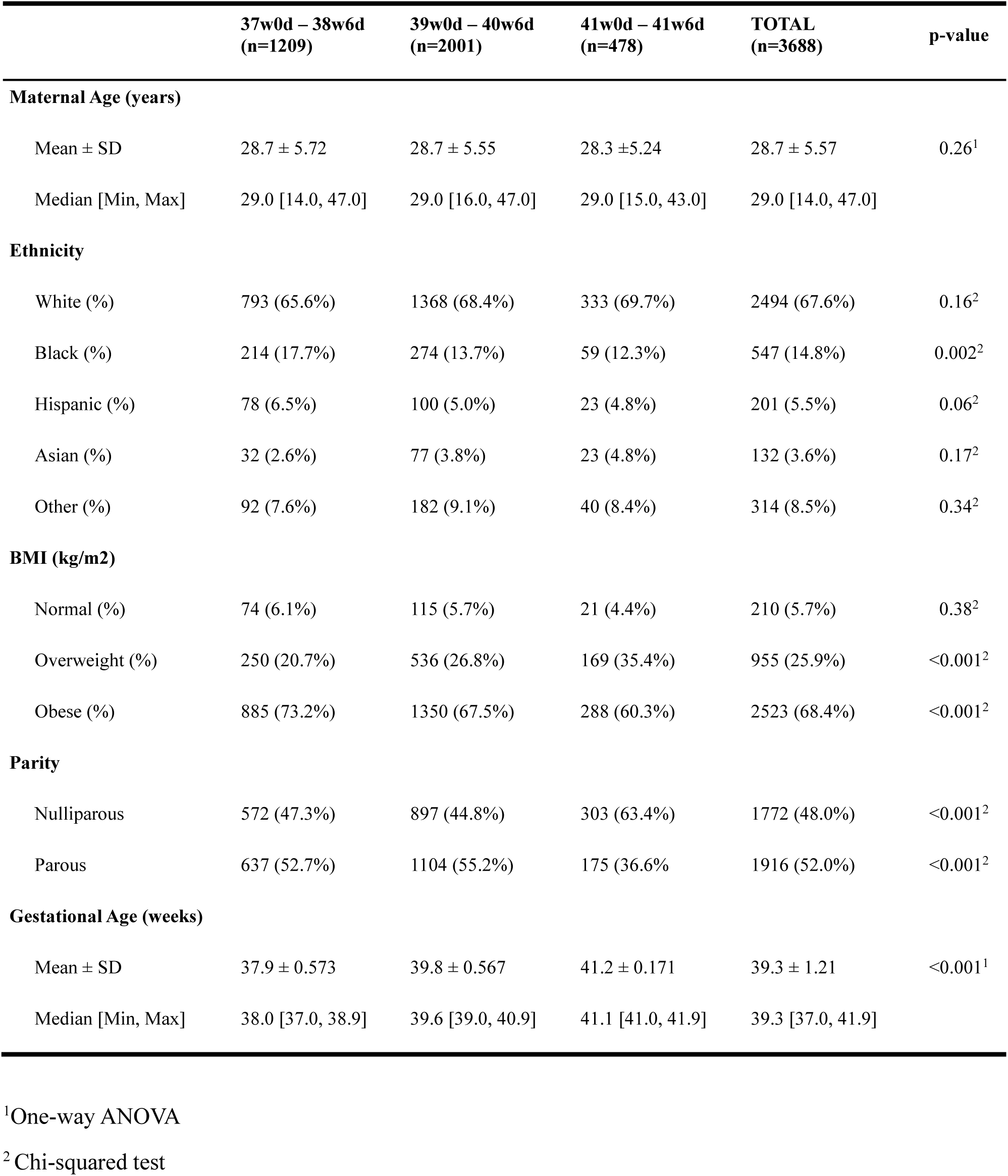
Population statistics of study participants comparing gestational age groups.

|  | 37w0d – 38w6d<br>(n=1209) | 39w0d – 40w6d<br>(n=2001) | 41w0d – 41w6d<br>(n=478) | TOTAL<br>(n=3688) | p-value |
| --- | --- | --- | --- | --- | --- |
| **Maternal Age (years)** |  |  |  |  |  |
| Mean ± SD | 28.7 ± 5.72 | 28.7 ± 5.55 | 28.3 ± 5.24 | 28.7 ± 5.57 | 0.26<sup>1</sup> |
| Median [Min, Max] | 29.0 [14.0, 47.0] | 29.0 [16.0, 47.0] | 29.0 [15.0, 43.0] | 29.0 [14.0, 47.0] |  |
| **Ethnicity** |  |  |  |  |  |
| White (%) | 793 (65.6%) | 1368 (68.4%) | 333 (69.7%) | 2494 (67.6%) | 0.16<sup>2</sup> |
| Black (%) | 214 (17.7%) | 274 (13.7%) | 59 (12.3%) | 547 (14.8%) | 0.002<sup>2</sup> |
| Hispanic (%) | 78 (6.5%) | 100 (5.0%) | 23 (4.8%) | 201 (5.5%) | 0.06<sup>2</sup> |
| Asian (%) | 32 (2.6%) | 77 (3.8%) | 23 (4.8%) | 132 (3.6%) | 0.17<sup>2</sup> |
| Other (%) | 92 (7.6%) | 182 (9.1%) | 40 (8.4%) | 314 (8.5%) | 0.34<sup>2</sup> |
| **BMI (kg/m2)** |  |  |  |  |  |
| Normal (%) | 74 (6.1%) | 115 (5.7%) | 21 (4.4%) | 210 (5.7%) | 0.38<sup>2</sup> |
| Overweight (%) | 250 (20.7%) | 536 (26.8%) | 169 (35.4%) | 955 (25.9%) | <0.001<sup>2</sup> |
| Obese (%) | 885 (73.2%) | 1350 (67.5%) | 288 (60.3%) | 2523 (68.4%) | <0.001<sup>2</sup> |
| **Parity** |  |  |  |  |  |
| Nulliparous | 572 (47.3%) | 897 (44.8%) | 303 (63.4%) | 1772 (48.0%) | <0.001<sup>2</sup> |
| Parous | 637 (52.7%) | 1104 (55.2%) | 175 (36.6%) | 1916 (52.0%) | <0.001<sup>2</sup> |
| **Gestational Age (weeks)** |  |  |  |  |  |
| Mean ± SD | 37.9 ± 0.573 | 39.8 ± 0.567 | 41.2 ± 0.171 | 39.3 ± 1.21 | <0.001<sup>1</sup> |
| Median [Min, Max] | 38.0 [37.0, 38.9] | 39.6 [39.0, 40.9] | 41.1 [41.0, 41.9] | 39.3 [37.0, 41.9] |  |

<sup>1</sup>One-way ANOVA
<sup>2</sup>Chi-squared test

### Data carpentry and visualization

Data carpentry and visualizations were performed under R environment (version 4.0.5). The *readxl* (version 1.3.1) package was used for loading the excel file format. Visualizations were prepared using the *ggplot2* (version 3.3.3) library and colors from *RColorBrewer* (version 1.1.2). Data carpentry was performed using base R*, dplyr* (version 1.0.6), and *tidyr* (version 1.1.3) packages. Statistical tests including ANOVA, Kruskal-wallis, and t-test were executed using *rstatix* (version 0.7.0).

### Circadian rhythm analysis

The rhythmic component of TOI was identified using the Lomb-Scargle method for circadian rhythm^18,19^ analysis and plotting a periodogram. The analysis was performed by separating TOI in one-hour bins for a total of 23 periods and α = 0.01 for identifying statistical significance. The specific peaks for identifying TOI responsible for greatest variation in the data and normalized power for assessing strength of the rhythm were analyzed.

### Multivariate analysis

PCA was performed with the *factoextra (version 1.0.7)* and *FactoMineR (version 2.4)* packages and used to identify key variables related to labor duration and failed induction of labor. Labor duration was binned by less than 12 hours, between 12 and 24 hours, and greater than 24 hours.

### Time-to-event analysis

Time-to-event analysis of total induction of labor (IOL) duration was performed using a standard survival analysis and Cox Proportional Hazard (CPH) analysis with the *survival* (version 3.2.11) and *survminer* (version 0.4.9) packages. Delivery was considered the event and hazard ratio (HR) is reported with 95% confidence interval (CI) as HR [95% CI]. All data was used as the reference for comparing each bin versus the rest of the sample.

## Results

### Baseline population characteristics

We did not detect significant differences in maternal age (p=0.26; One-way ANOVA) between GA groups (Table 1). We observed significant differences in ethnicity (p=0.004; Chi-Square test), BMI (p=<0.001; Chi-Square test), Parity (p=<0.001; Chi-Square test) and Gestational Age (p=<0.001; One-way ANOVA) among the groups (Table 1). In addition, we found differences in the proportion of women in each GA group according to ethnicity. A significantly higher proportion of the Black women were induced between 37w0d – 38w6d compared to 39w0d – 40w6d and 41w0d – 41w6d (17.7% vs. 13.7% and 12.3%, p=0.002). There were no significant differences between GA groups in White (p=0.158; Chi-squared test), Hispanic (p=0.06), Asian (p=0.17), or Other Ethnicity (p=0.34). Labor induction was significantly different across GA groups in overweight (20.7% for 37w0d – 38w6d, 26.8% for 39w0d – 40w6d, 35.4% for 41w0d – 41w6d, p<0.001; Chi-squared test) and obese (73.2% for 37w0d – 38w6d, 67.5% for 39w0d – 40w6d, 60.3% for 41w0d – 41w6d, p=0.38 pregnancies, as compared to normal weight (p=0.38) pregnancies.

IOL duration was significantly longer in women having a cesarian section as compared to vaginal delivery (p<0.001, Welch’s two-sample t-test; Supplementary Figure 1A). Compared to the rest of the induction cohort, women with IOL duration less than 12 hours resulted in a significantly reduced the odds ratio (OR) of cesarean section (0.350 [0.290, 0.419], p<0.001; Supplementary Figure 1B). Women with IOL duration beyond 24 hours had a significantly increased OR of cesarean section (2.999 [2.553, 3.524], p<0.001). Women with IOL duration falling between 12 and 24 hours showed no significant differences (p=0.29).

### Labor duration is shortest with induction during early morning hours

Investigating the average IOL duration, regardless of induction start method, at each TOI revealed shorter durations during early morning hours. Average induction duration was shortest with TOI at 5:00, resulting in an average of 14.6 hours (Figure 1A). IOL duration gradually increased throughout the day, peaking with TOI at 23:00 hours, resulting in an average duration of 20.6 hours. Total IOL duration was significantly different between these TOI (p=0.001, Kruskal-Wallis test). Results of pairwise post hoc analysis by Dunn’s test are provided in Supplemental Table 1. Further stratification by induction start method revealed additional trends in the IOL duration warranting further investigation using larger datasets for each induction method (Supplemental Figure 2). In this study, the entire dataset including each induction start method was retained in the downstream analysis to maintain statistical power.

**Figure 1.**
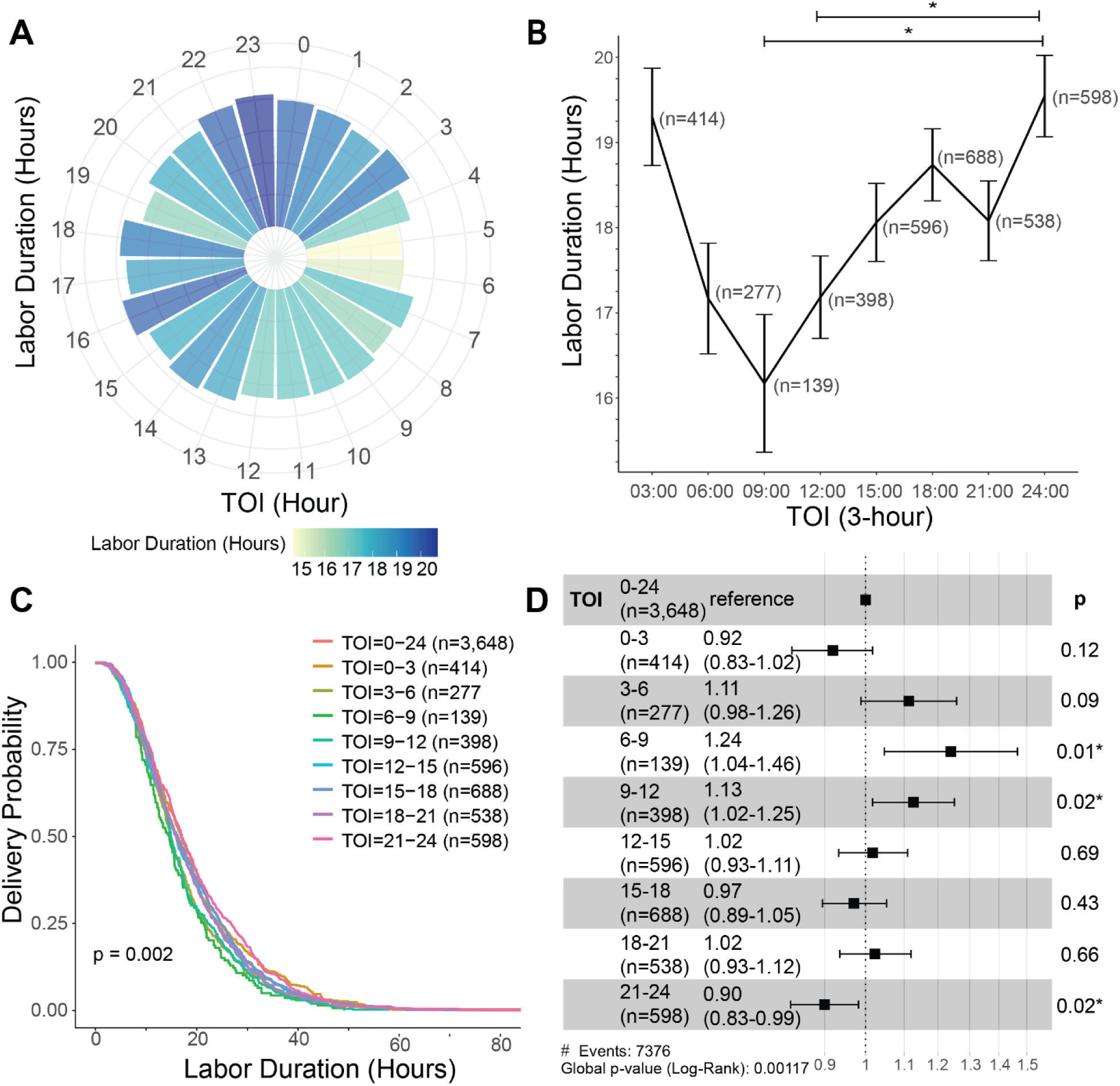
Time of Induction (TOI) defines labor duration in term women. The average labor duration for each TOI in 1-hour bins (panel A) is shown as a clock-like pattern from 0:00 to 23:00 hours. Labor duration is significantly different between TOI bins (p=0.001, Kruksal-Wallis) and durations are indicated by color gradient from shorter durations in yellow, over green, to longer durations in blue. *Post-hoc* analysis using Dunn’s test can be found in Supplementary Table 1. Further aggregating into 3-hour bins (panel B) was implemented to maintain statistical power in downstream analysis and shows significant differences between TOI bins (p=0.001, one-way ANOVA). Time-to-event analysis was applied using Kaplan-Meier curves (panel C) and Cox Proportional Hazard (CPH) (panel D) for each TOI in 3-hour bins. Delivery probability was defined as probability of giving birth and the event in hazard ratio (HR) defined as giving birth. Higher HR values denote a higher probability of reaching birth in that TOI bin compared to any other time of day (0:00-23:00) as a reference. Significant differences, p-values, and n/bin are indicated in panels B-D.

Subsequently, we observed differences in labor progression contributing to the significant difference in total IOL duration across TOI groups (p=0.01, Kruskal-Wallis test, Supplemental Figure 3). There were no significant differences in duration from cervical dilation of 6 cm to second stage of labor (p=0.62), or from second stage of labor to delivery (p=0.45).

Next, we aimed to determine whether IOL duration was rhythmically dependent on TOI. Lomb-Scargle periodogram analysis of periodicity of labor duration in the entire induced population identified a significant circadian rhythm (p=0.02, Supplementary Figure 4A). Moreover, since BMI is known to impact labor duration and Pitocin efficacy^20^, the data was further stratified into normal weight, overweight, and obese according to BMI. We did not identify a significant rhythmicity in normal weight women (p=0.94, Supplementary Figure 4B) or overweight women (p=0.95, Supplementary Figure 4C), but we did see significant rhythmicity in the obese population (p=0.002, Supplementary Figure 4D).

We then binned the data in 3-hour increments to increase the number of samples per bin and observed significant differences in IOL duration depending on TOI (Figure 1B, p=0.001, one-way ANOVA). Women induced between 6:00-9:00 (adjusted p=0.03) or 9:00-12:00 (adjusted p=0.03) had significantly shorter labor duration than women induced from 21:00-24:00. Inducing labor between 6:00-9:00 or 9:00- 12:00 resulted in an average labor duration of 16.2 and 17.1 hours compared to 19.5 hours at the highest point with induction between 21:00-24:00.

Time-to-birth analysis was performed using a standard survival analysis to calculate the probability of delivery. Delivery probability was defined as the probability that a pregnant woman gives birth from the time origin to a specified future time. We identified significant differences in the probability of delivery based on TOI (Figure 1C, p=0.002). We identified a significantly increased chance of birth (Figure 1D) with induction between 6:00-9:00 hours (1.24 [1.04-1.45], p=0.014) and 9:00-12:00 hours (1.13 [1.02- 1.25], p=0.02), demonstrating an increased likelihood to reach delivery, irrespective of delivery method, with induction during this timeframe. On the contrary, labor induction between 21:00-24:00 hours resulted in a significantly decreased delivery rate (0.90 [0.83-0.99], p=0.023) suggesting late evening inductions may increase time to delivery, which is a risk factor for cesarean sections (Supplemental Figure 1A).

### Time-of-day of labor induction is not associated with increased rate of cesarean section or infant outcomes

To determine whether inducing labor at a particular time of the day would be an associated risk for labor outcome, we studied the relationship between TOI and delivery methods, NICU admissions, and resulting infant outcomes. We found no significant differences looking at delivery method (Supplementary Figure 5A) based on TOI (p=0.75, chi-squared test), as well as no differences in OR for cesarean section delivery (Supplementary Figure 5C; Supplementary Table 2) for any of the TOI. Additionally, we found no significant differences in NICU admission (Supplementary Figure 5B, p=0.66, Chi-square) or OR for being admitted to the NICU (Supplementary Figure 5D; Supplementary Table 2) across TOI. Likewise, we found no significant difference in infant diagnoses based on TOI for sepsis (p=0.61, Supplementary Figure 6A), respiratory distress (p=0.65, Supplementary Figure 6B), transient tachypnea (p=0.51, Supplementary Figure 6C), hypoglycemia (p=0.26, Supplementary Figure 6D), feeding difficulties (p=0.34, Supplementary Figure 6E), or jaundice (p=0.83, Supplementary Figure 6F).

### Maternal factors condition effect of TOI on labor duration

Principal Component Analysis (PCA) showed clusters according to IOL duration and maternal factors. Based in two-dimensions plotting (Figure 2A), there is a continuous separation in which pregnant women with IOL duration beyond 24 hours (red dots) separate from the other groups followed by women who labor between 12 and 24 hours (yellow dots), and less than 12 hours (blue dots). Graph of variables (Figure 2B) revealed pre-pregnancy weight and BMI as the top variables contributing to this separation in Dimension 2 (Dim 2). Other top contributors in Dim 2 of the PCA variable plot include labor induction variables, such as the number of doses, minutes from induction to delivery, and time from induction to dilation of 6 cm. Top contributors in Dim 1 include maternal variables, such as gravida, para and term births. Based on the PCA analysis, the data was controlled for the top three variables which conditioned labor duration as a function of TOI: BMI, parity, and GA. Pregnant women with a normal range BMI experienced the shortest interval to delivery with induction from 0:00-3:00 (12.01 ± 6.41 hours) and peaking with the longest IOL when induced between 6:00-9:00 (17.15 ± 4.36 hours, Figure 3A). This trend shifted to later in the day in overweight pregnant women, resulting in the shortest IOL duration with TOI between 3:00-6:00 (15.33 ± 10.43 hours), and peaking with the longest interval to delivery with TOI between 12:00-15:00 (18.24 ± 12.44 hours). Obese women experienced the shortest interval to delivery with TOI between 6:00-9:00 (16.42 ± 9.52 hours), peaking at 15:00-18:00 (20.15 ± 11.75 hours) and again from 21:00-24:00 (21.07 ± 12.02 hours). Two-way ANOVA revealed significant differences with 3- hour induction bins (p=0.001) and BMI (p<0.001), but interactions between the variables were not significant (p=0.07).

**Figure 2.**
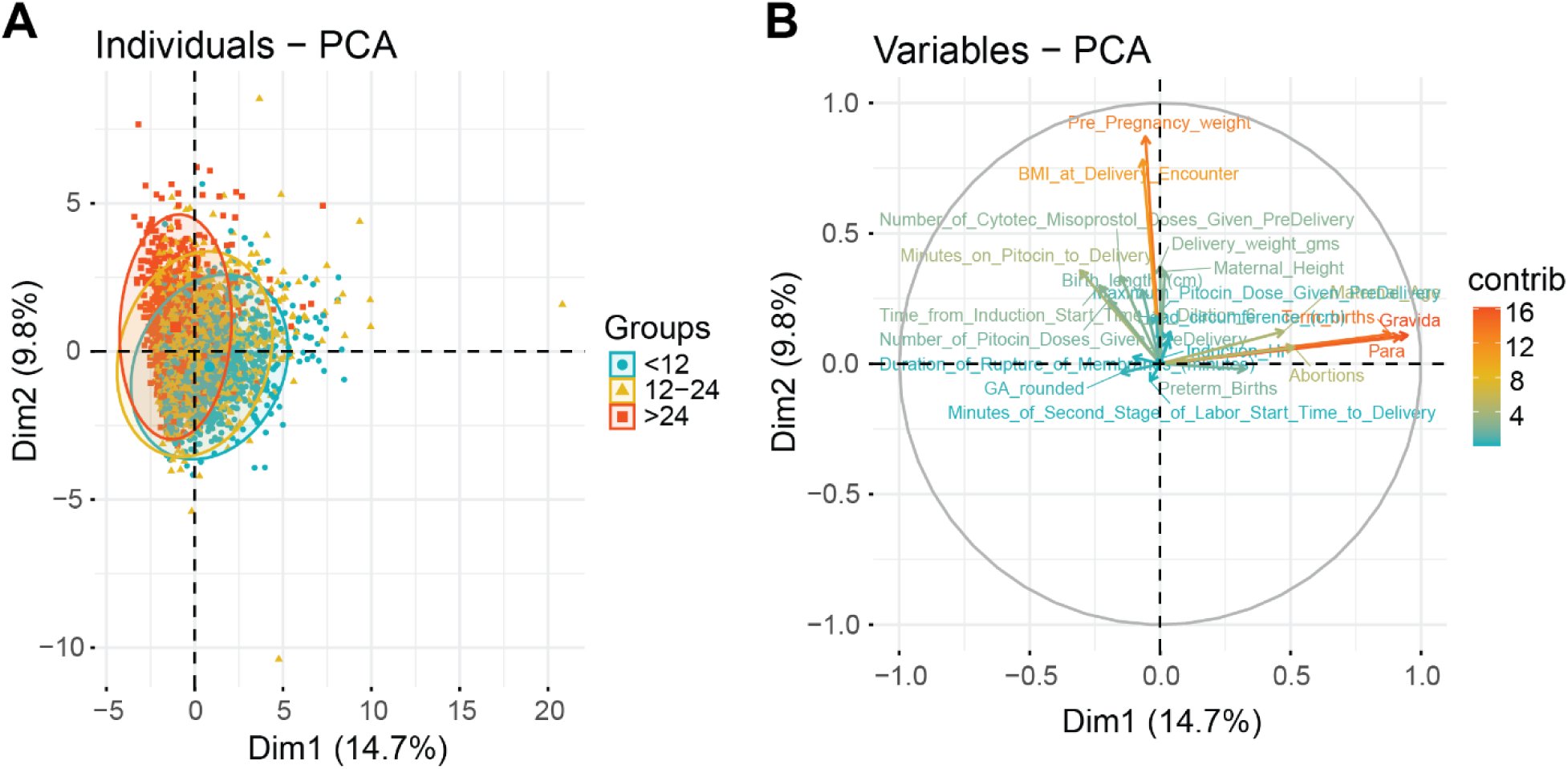
Multivariate analysis revealed clusters according to labor duration. Principle Component Analysis (PCA) was performed, and the plot of individuals (panel A) was grouped by labor duration greater than 24 hours (red), between 12 and 24 hours (yellow), and shorter than 12 hours (blue). Graphic of variables was plotted with the direction and contribution for each variable in the PCA analysis (panel B). Positively and negatively correlated variables point to the same or opposite side of the plot, respectively. Strength of the contribution is indicated by the length of the arrow and color gradient with longer arrows and red color indicating higher contributions, while lower contributions are shorter in length and indicated by the color blue. Women who labor beyond 24 hours begin to separate from the other groups with top contributors of labor duration identified as body mass index (BMI), pre-pregnancy weight, gravida, parity, and term births.

**Figure 3.**
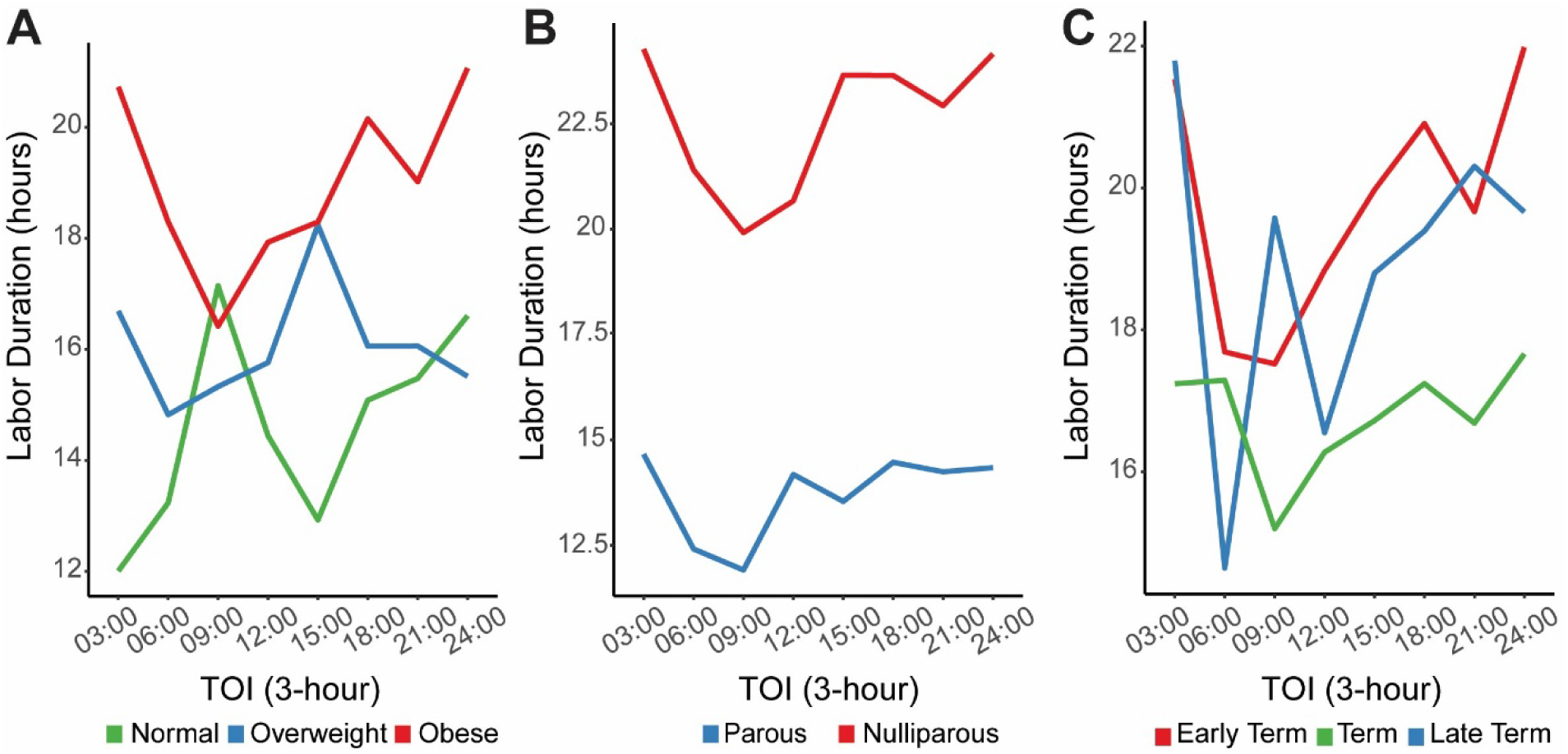
Effect of induction time-of-day on labor duration is conditioned by body mass index (BMI), parity, and gestational age (GA). Multivariate effect of time of induction (TOI) on labor duration with respect to maternal BMI (panel A), parity (panel B), and GA grouping at term (panel C). BMI group (p=0.07, two-way ANOVA) is indicated by color for normal weight (green) (n=205), overweight (blue) (n=941), and obese women (red) (n=2,502). Parity (p=0.21, two-way ANOVA) is indicated by color with nulliparous women (n=1,735) defined as P0 and red in color, while parous women (n=1,913) are defined as P1+ and indicated by blue. GA groups at term (p=0.45, two-way ANOVA) are defined as early term (red) (n=1,193) from 37 weeks 0 days to 38 weeks 6 days, term (green) (n=1,983) from 39 weeks 0 days to 40 weeks 6 days, and late term (blue) (n=472) from 41 weeks 0 days to 41 weeks 6 days. Women who delivered before 37 weeks or after 42 weeks were not included in this dataset.

In agreement with the literature^21^, nulliparous women experienced a longer labor duration compared to parous women regardless of TOI (Figure 3B). Interestingly, the average labor duration was shortest with induction during morning hours regardless of parity (Figure 3B). Both nulliparous and parous women experienced the shortest IOL duration with induction between 6:00-9:00 (19.91 ± 10.04 hours). Two-way ANOVA revealed significant differences with 3-hour induction bins (p<0.001) and parity (p<0.001), but interactions between the variables were not significant (p=0.21).

To determine the impact of GA, data was stratified by early term, term, and late term (Figure 3C). IOL duration of term deliveries was less variable than early term and late term, with early term deliveries displaying a longer average labor duration compared to the other groups. Both early term and term deliveries displayed a similar trend with shorter IOL duration with morning induction and gradually increasing throughout the day. Late-term deliveries revealed variation in labor duration across induction times with no significant differences. Two-way ANOVA revealed significant differences with 3-hour induction bins (p=0.001) and GA groups (p<0.001), but interactions between the variables were not significant (p=0.45).

To determine the effect of maternal factors on labor duration as a function of induction time, we repeated the time-to-event analysis stratified by BMI and parity (Figure 4). We found significant differences in delivery probability based on TOI in the nulliparous obese (p=0.02) and parous obese populations (p=0.03). Nulliparous obese women hade a significantly increased chance of giving birth with induction between 6:00-9:00 (HR [95% CI] = 1.37 [1.03, 1.80], p=0.03) and between 9:00-12:00 (HR [95% CI] = 1.22 [1.01, 1.50], p=0.04). Furthermore, parous obese women had a significantly increased chance of giving birth with induction between 3:00-6:00 (HR [95% CI] = 1.25 [1.00, 1.50], p=0.05) and 6:00-9:00 (HR [95% CI] = 1.50 [1.12, 2.0], p=0.007)

**Figure 4.**
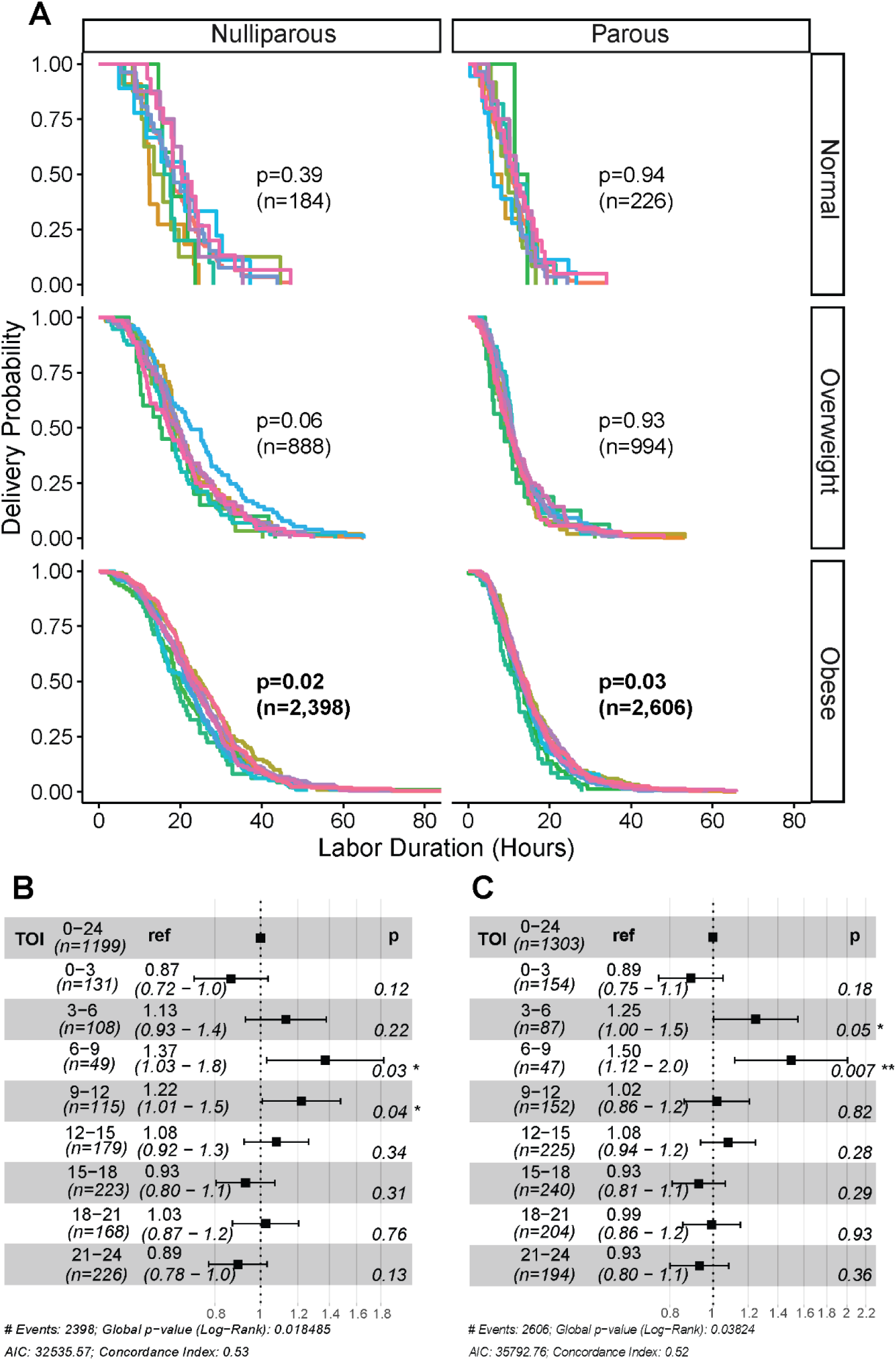
Time-to-event analysis of TOI identifies differences in delivery probability as a function of body mass index (BMI) and parity. Time-to-event analysis of labor duration across time of induction (TOI) groups in 3-hour bins was stratified by BMI and parity. Kaplan-Meier curves were produced for each combination of BMI and parity (panel A) and the resulting p-values indicated in the plots. Delivery probability was defined as probability of giving birth. Nulliparous obese and parous obese groups with significant differences in delivery probability across TOI were selected for Cox Proportional Hazard (CPH) analysis (panels B and C). The event in hazard ratio (HR) was defined as giving birth. Higher HR values denote a higher probability of reaching birth in that TOI bin compared to any other time of day (0:00-23:00) as a reference. Significance was set at α=0.05 with p-values and n/bin indicated in each panel.

## Discussion

Here, we describe for the first time that labor induction has a circadian rhythm, and TOI influences the resulting IOL duration. Importantly, we show that the optimal TOI is conditioned by maternal phenotype and consideration of induction time-of-day may be most beneficial for specific at-risk populations.

Comparative analysis of maternal baseline characteristics across GA groups revealed small, yet significant differences in ethnicity, BMI, and Parity. Such differences have been observed in other studies^22–24^ and highlight the consistent impact of maternal factors in induction success and delivery outcomes. Importantly, we confirm that BMI and parity are significant contributing factors for labor duration^25^. Stratifying these variables uncovered different TOI patterns for these groups, where obese and nulliparous women were most susceptible to experiencing a longer labor duration following induction of labor. However, nulliparous obese and parous obese women had significantly different labor durations across TOI groups, demonstrating these populations stand to benefit the most from planned induction during the optimal window of time.

Our identification of TOI as defining labor duration provides a novel and cost-effective strategy to improve the management of women in labor. Previous studies on the management of laboring women have primarily focused on hospital staffing and shifts^26–28^, and not considered TOI from the standpoint of the maternal circadian system. Studies who have addressed TOI as a variable found no differences in the average duration of labor between morning and evening induction^29^, indicating the need for a more granulated analysis of the data to identify the optimal TOI. In agreement with our data, others have reported no differences in maternal or neonatal outcomes with morning versus evening induction^30^, supporting the safety of aligning the TOI to maternal characteristics to reduce labor duration. We did not identify a reduced rate of cesarian section or NICU admissions as a function of TOI. This lack of significance might be due to insufficient power in our dataset with TOI in 3-hour bins. In agreement with the literature, we found no significant differences in delivery method or infant outcomes across TOI groups. Hence, inducing expectant mothers at the optimal time-of-day based on their characteristics has the potential to reduce labor duration without an added risk of delivering by cesarian section, NICU admission, or resulting infant diagnoses. This supports the safety of considering TOI to improve management of women who are induced without negative maternal or fetal consequences associated.

In conclusion, this study provides strong evidence for the use of TOI as a novel consideration for labor management. We propose careful consideration of TOI. In particular, nulliparous and parous women with increased BMI, who are most prone to increased labor duration, could benefit the most from inducing labor in the early morning between 3:00-9:00, a time window that is associated with shorter labor duration across all groups.

## Data Availability

All data produced in the present work are contained in the manuscript.

## FUNDING SOURCES

This work was funded by the USDA National Institute of Food and Agriculture Hatch project (MICL1018024) to HMH, the NIH/National Institute of Environmental Health Sciences project (R01ES035691) to HMH, and a sub-award from the Michigan Diabetes Research Center through the National Institute of Diabetes and Digestive and Kidney Diseases (P30DK020572).

## SUPPLEMENTARY MATERIAL

**Supplementary Figure 1.**
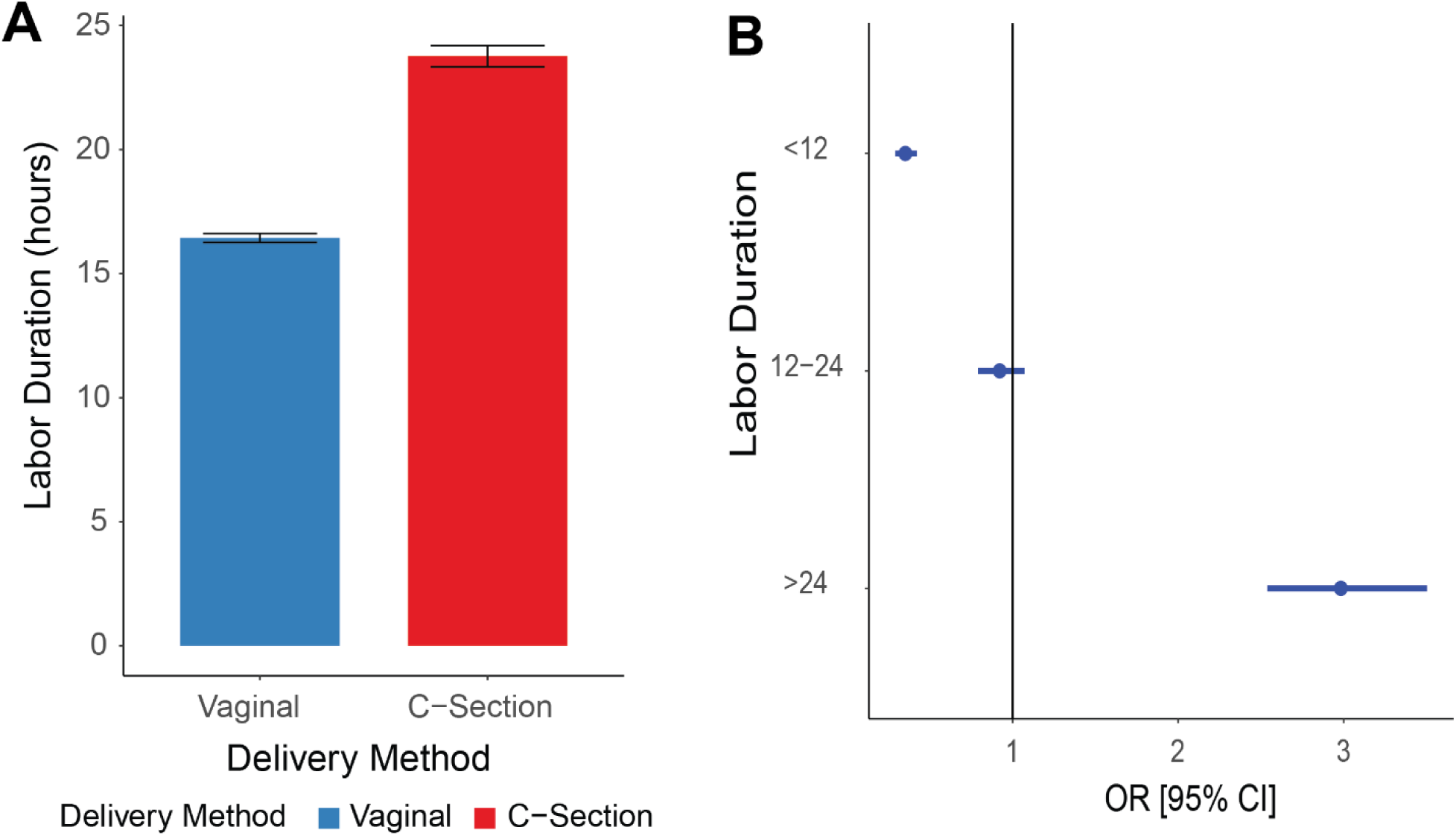
Longer labor duration increases the odds of delivering by cesarean section. Average labor duration in hours for vaginal (n=2,714) and cesarean section (n=934) deliveries (panel A) and odds ratio (OR) of delivering by cesarean section across labor duration bins (panel B). The point in panel B denotes OR with the 95% confidence interval (CI) indicated by the blue lines. OR=1 is shown with the vertical black line. Labor duration less than 12 hours was significantly different from the other groups OR [95% CI] = (0.350 [0.290, 0.419], p<0.001, n=1,215). Labor duration greater than 24 hours was also significantly different from the other groups (2.999 [2.553, 3.524], p<0.001, n=922). Labor duration between 12 and 24 hours was not significantly different (p=0.29, n=1,511).

**Supplementary Figure 2.**
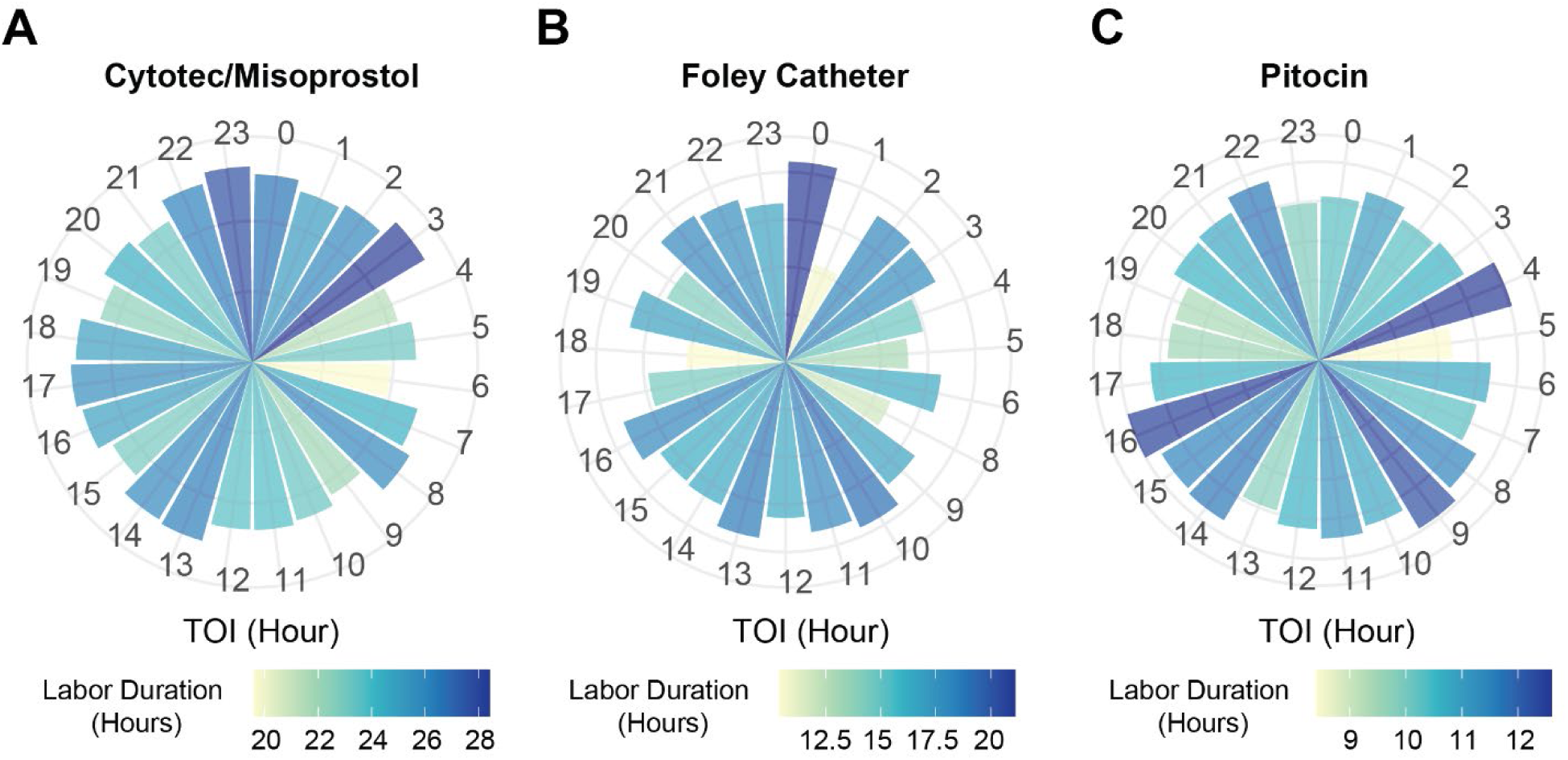
Effect of induction time stratified by induction start method. Average labor duration for each time of induction (TOI) in 1-hour bins for each induction start method of cytotec/misoprostol (panel A) (n=2,049; p=0.08, Kruskal-Wallis), foley catheter (panel B) (n=288; p=0.29, Kruskal-Wallis), and Pitocin (panel C) (n=1,311; p=0.04, Kruskal-Wallis). TOI is in a clock-like pattern from 0:00 to 23:00 hours. Labor duration is indicated by color gradient from shorter durations in yellow, over green, to longer durations in blue.

**Supplementary Figure 3.**
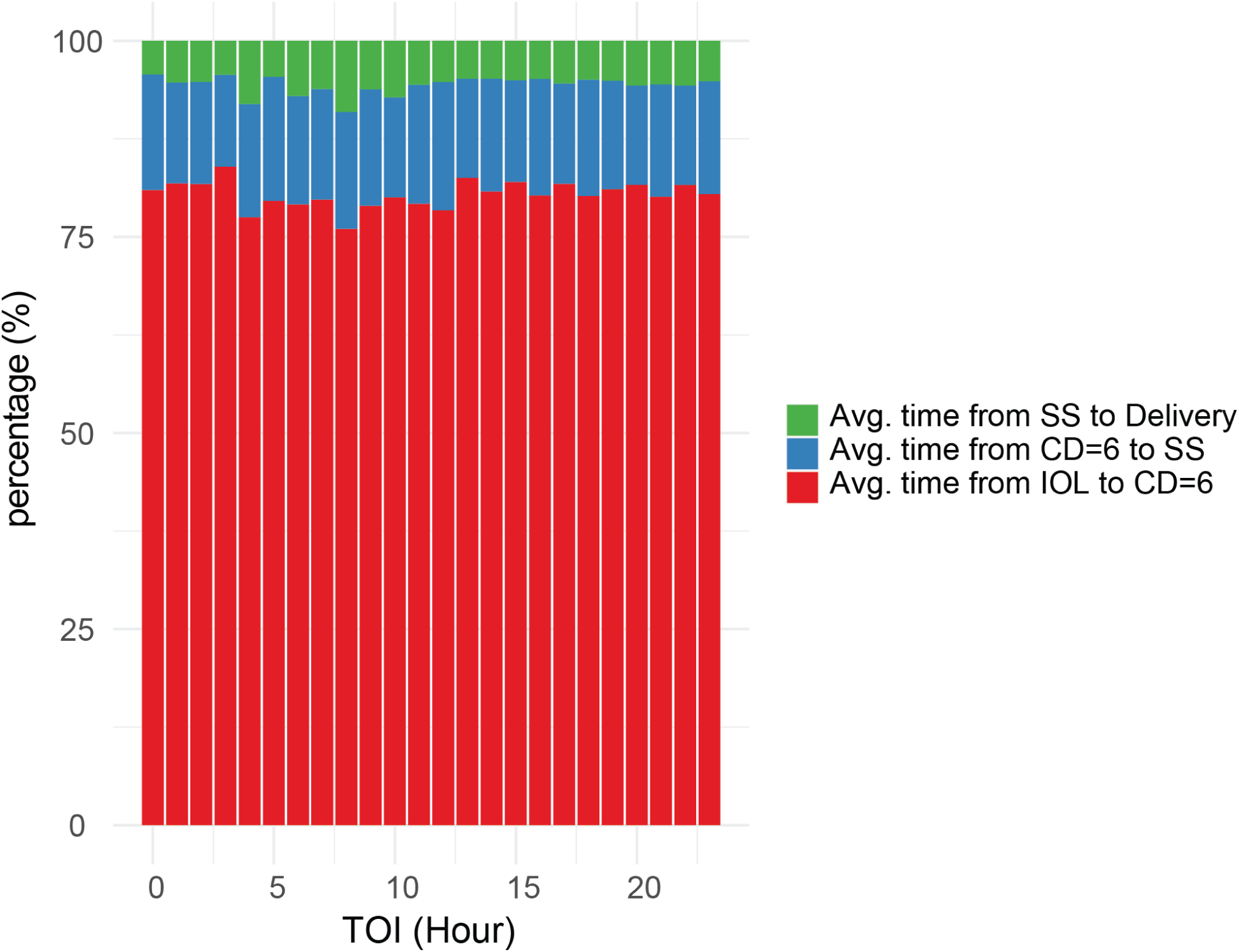
Labor progression across induction times. Percentages of total time in each stage of labor across 1-hour time of induction (TOI) bins. Stages of labor were defined as time from induction of labor (IOL) to cervical dilation (CD) of 6cm (red) (p=0.01, one-way ANOVA), time from cervical dilation of 6cm to second stage (SS) of labor (blue) (p=0.62, one-way ANOVA), and time from second stage of labor to delivery (green) (p=0.45, one-way ANOVA).

**Supplementary Figure 4.**
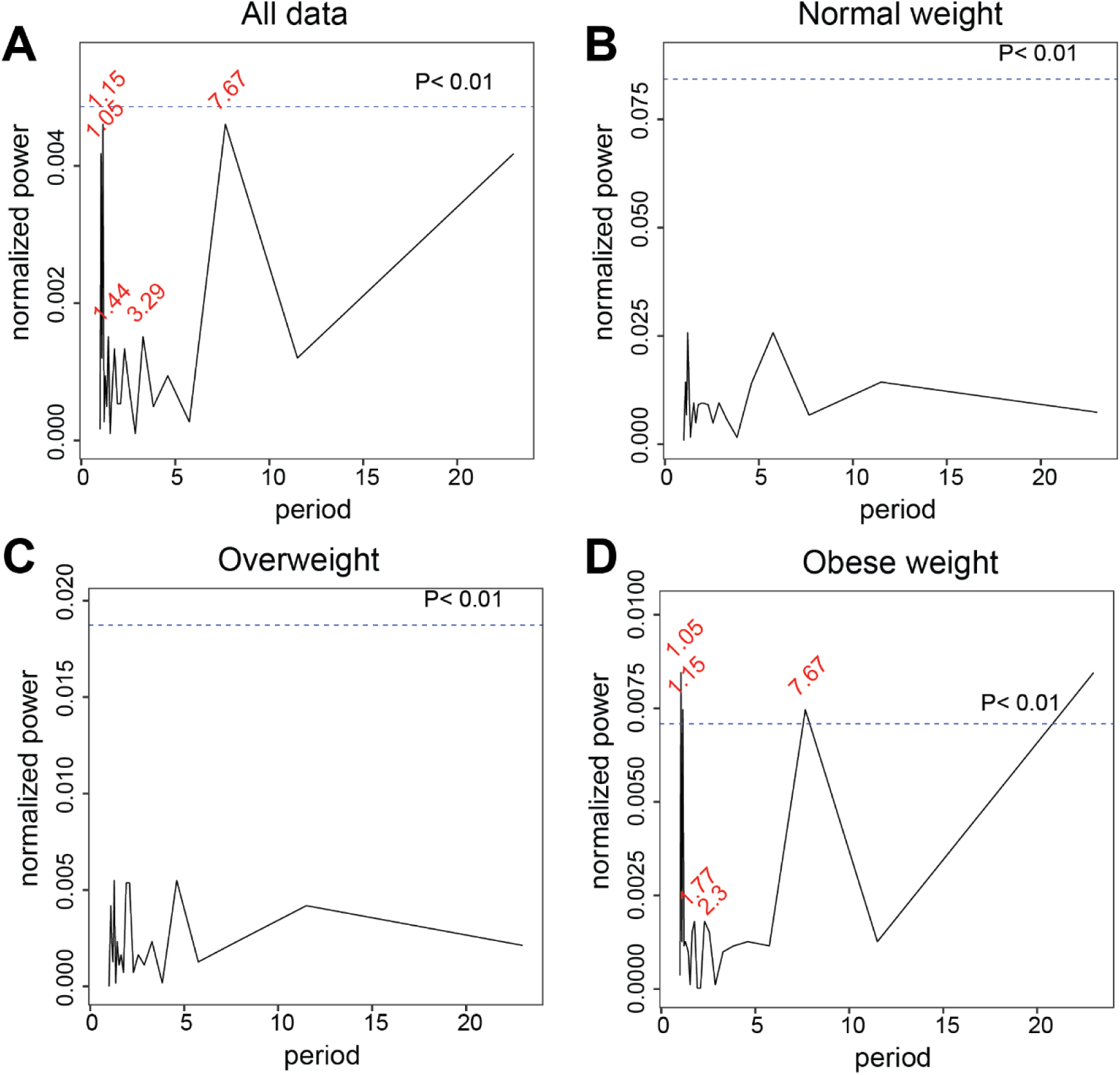
TOI impacts labor duration in a circadian manner and is influenced by BMI. The circadian rhythm analysis of time of induction (TOI) is shown using Lomb-Scargle Periodograms. The analysis was performed by separating TOI in one-hour bins for a total of 23 periods (x-axis) and α = 0.01 for identifying statistical significance, indicated by the dashed horizontal line. Specific peaks are identified in red for TOI bins responsible for the greatest variation in the data. Normalized power measures strength of the rhythm. There was a significant circadian rhythm of labor duration across the entire dataset (panel A) as a result of induction time, identified by significant peaks at 1.15 and 7.67 hours (p=0.0156, n=3,648). Separating by BMI reveals no significant peaks in normal weight (panel B) (p=0.94, n=205) or overweight (panel C) (p=0.95, n=941) pregnancies. Obese women (panel D) have a significant circadian rhythm (p=0.002, n=2,502) with peaks at 1.05, 1.15, and 7.67 hours.

**Supplementary Figure 5.**
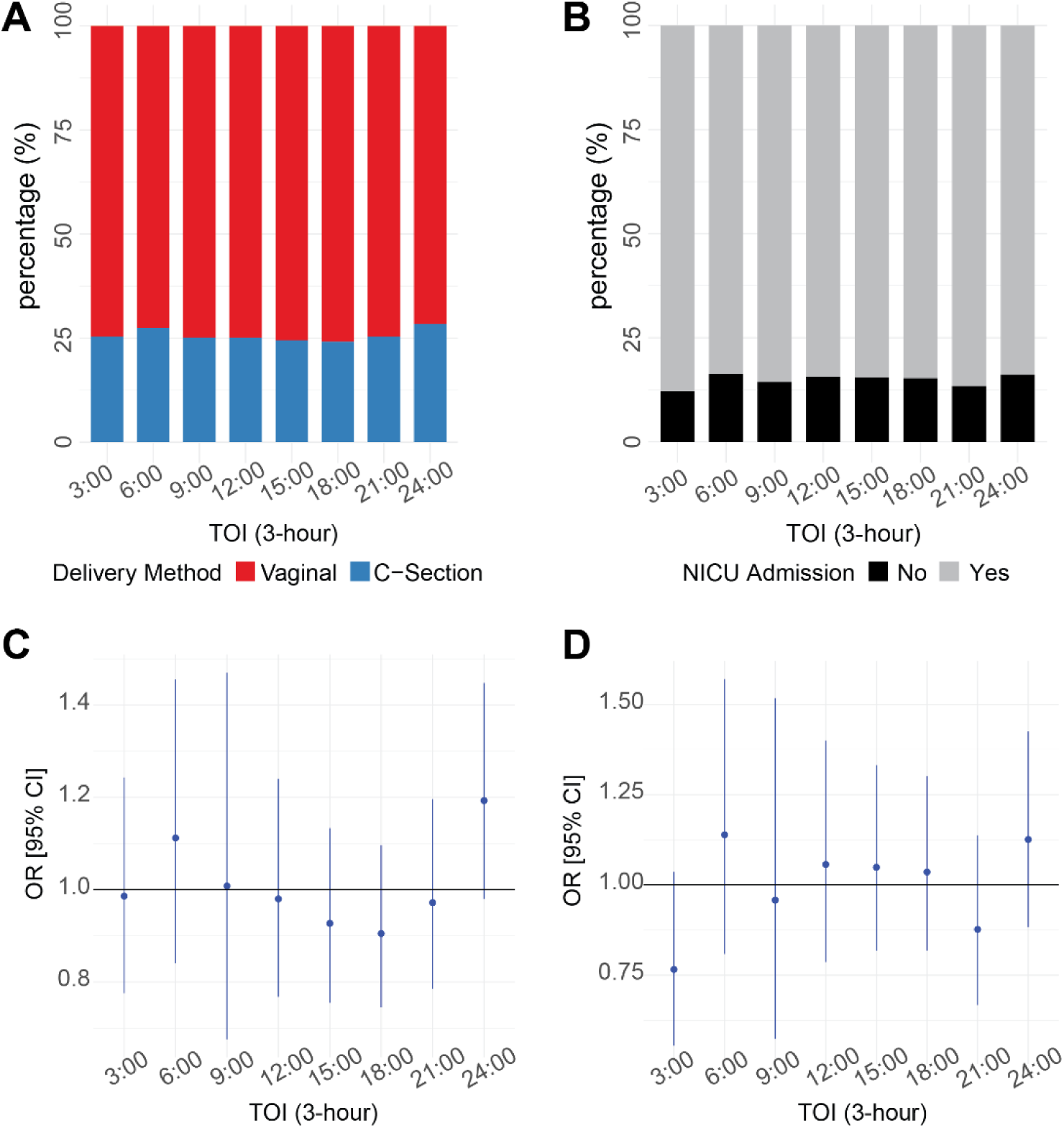
Odds ratio (OR) for delivery methods and NICU admissions as a function of the time of day of labor induction shows no significant differences. Shown are the percentage of vaginal (blue) (n=2,714) and cesarean sections (red) (n=934) for each time of induction (TOI) in 3-hour bins (panel A). The percentage of cesarean sections was highest with induction from 21:00-24:00 hours, accounting for 28.4% of deliveries at this timepoint. Differences in cesarean delivery method were not statistically significant (p=0.75, Chi-Square test). Panel B displays the percentage of NICU admissions for each TOI in 3-hour bins, also revealing the highest number of admissions with induction from 21:00- 24:00 hours, resulting in 16.1% of total deliveries at this timepoint. Differences in NICU admission were not statistically significant (n=542 admitted and n=3,106 not admitted; p=0.66, Chi-Square test). OR of delivering via cesarean section (panel C) and OR of being admitted to the NICU (panel D) were analyzed. OR of cesarean section and NICU admissions were not significantly different across TOI bins (refer to Supplementary Table 1 for individual p-values).

**Supplementary Figure 6.**
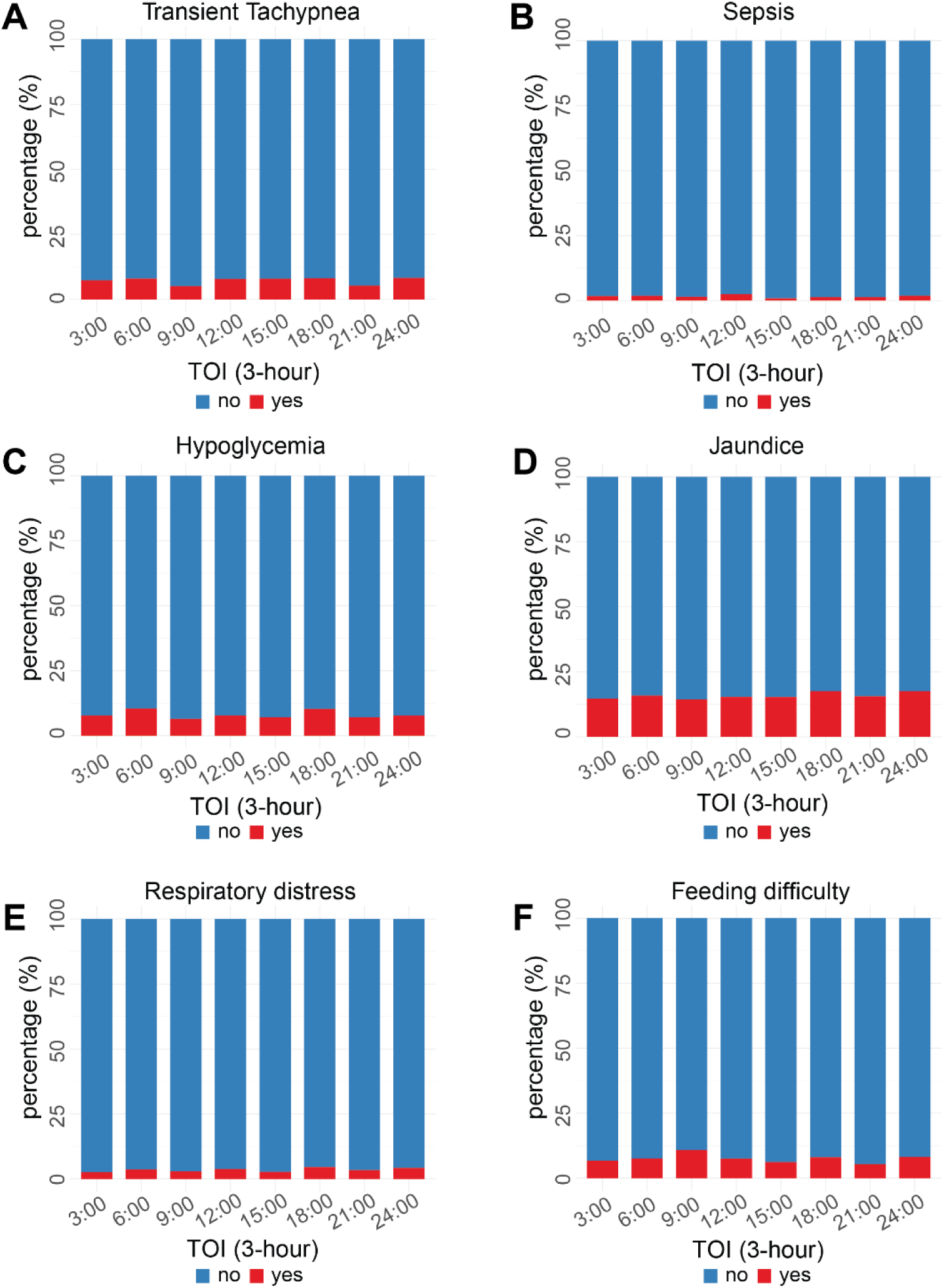
Time of induction (TOI) does not impact resulting infant diagnoses. Shown are the percentage of infant diagnoses for each TOI in 3-hour bins. Presence of the diagnosis is indicated by the color red, and absence of the diagnosis indicated by the color blue. Non-significant fluctuations are observed for sepsis (panel A) (p=0.61, Chi-Square test), respiratory distress (panel B) (p=0.65, Chi-Square test), transient tachypnea (panel C) (p=0.51, Chi-Square test), hypoglycemia (panel D) (p=0.25, Chi-Square test), feeding difficulties (panel E) (p=0.34, Chi-Square test), and jaundice (panel F) (p=0.83, Chi-Square test).

---

**Supplementary Table 1. Dunn Test Post-Hoc Analysis of time of induction (TOI) in 1-hour bins.\***

**Supplementary Table 2.**
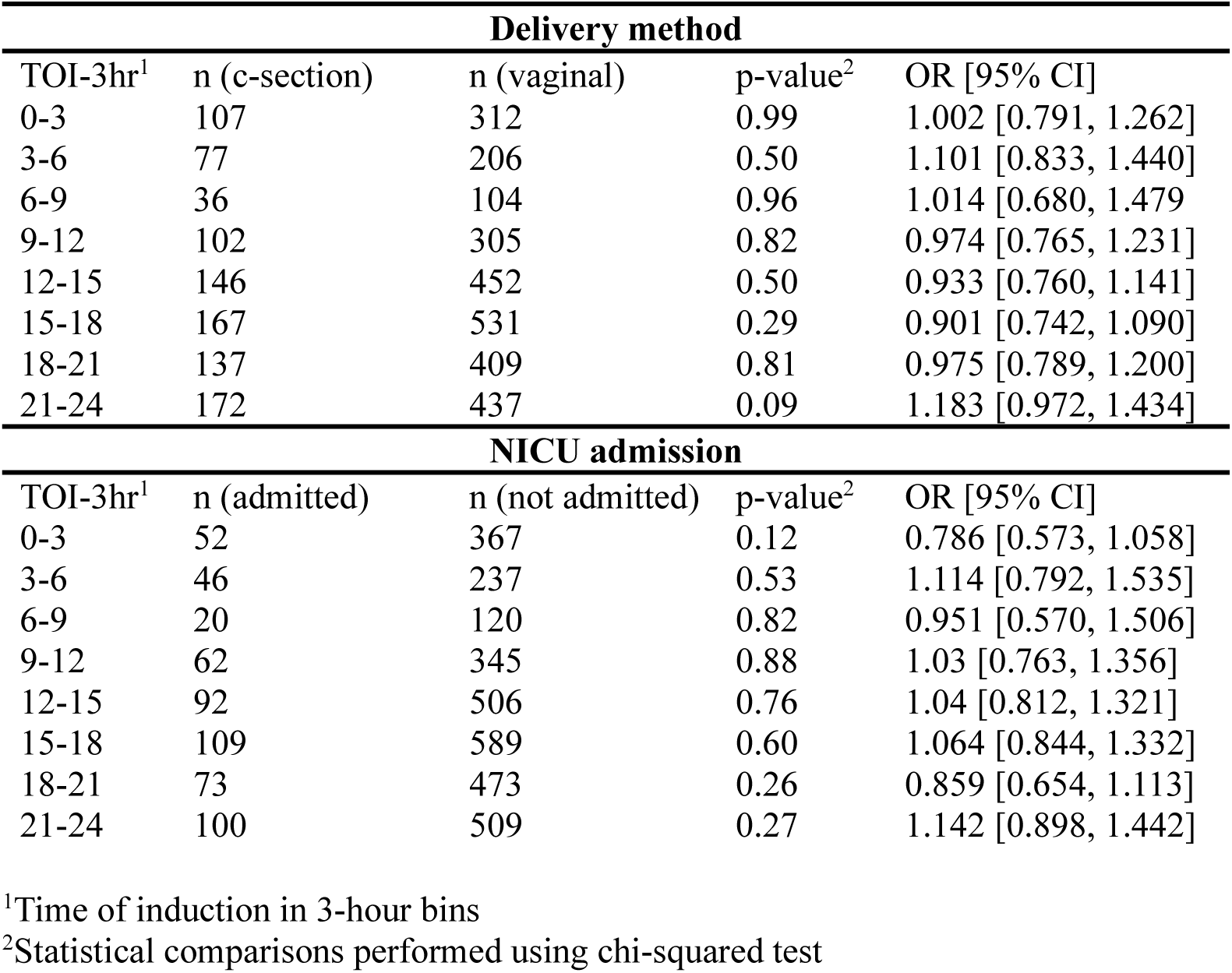
Odds ratios (OR) of delivery method and NICU admissions.

## Notes

### Competing Interest Statement

The authors have declared no competing interest.

### Author Declarations

This project is approved by the Institutional Review Board of the Michigan State University under Study ID: 0007199.

