## Supplemental Table 2 for "LABOR DURATION IS DEFINED BY THE TIME OF DAY OF LABOR INDUCTION"

**Supplementary Table 2. Odds ratios (OR) of delivery method and NICU admissions.**

| **Delivery method** | | | | |
| --- | --- | --- | --- | --- |
| TOI-3hr^1^ | n (c-section) | n (vaginal) | p-value^2^ | OR [95% CI] |
| 0-3 | 107 | 312 | 0.99 | 1.002 [0.791, 1.262] |
| 3-6 | 77 | 206 | 0.50 | 1.101 [0.833, 1.440] |
| 6-9 | 36 | 104 | 0.96 | 1.014 [0.680, 1.479 |
| 9-12 | 102 | 305 | 0.82 | 0.974 [0.765, 1.231] |
| 12-15 | 146 | 452 | 0.50 | 0.933 [0.760, 1.141] |
| 15-18 | 167 | 531 | 0.29 | 0.901 [0.742, 1.090] |
| 18-21 | 137 | 409 | 0.81 | 0.975 [0.789, 1.200] |
| 21-24 | 172 | 437 | 0.09 | 1.183 [0.972, 1.434] |
| **NICU admission** | | | | |
| TOI-3hr^1^ | n (admitted) | n (not admitted) | p-value^2^ | OR [95% CI] |
| 0-3 | 52 | 367 | 0.12 | 0.786 [0.573, 1.058] |
| 3-6 | 46 | 237 | 0.53 | 1.114 [0.792, 1.535] |
| 6-9 | 20 | 120 | 0.82 | 0.951 [0.570, 1.506] |
| 9-12 | 62 | 345 | 0.88 | 1.03 [0.763, 1.356] |
| 12-15 | 92 | 506 | 0.76 | 1.04 [0.812, 1.321] |
| 15-18 | 109 | 589 | 0.60 | 1.064 [0.844, 1.332] |
| 18-21 | 73 | 473 | 0.26 | 0.859 [0.654, 1.113] |
| 21-24 | 100 | 509 | 0.27 | 1.142 [0.898, 1.442] |

^1^Time of induction in 3-hour bins

^2^Statistical comparisons performed using chi-squared test
